# A Multidomain Model for Dementia Classification using Harmonized LASI and LASI-DAD Data

**DOI:** 10.64898/2026.06.13.26354833

**Authors:** Shweta Anand, Krishna P. Miyapuram

**Author notes:** **Corresponding author** - Shweta Anand, Cognitive Science Alumnus, Indian Institute of Technology Gandhinagar, Palaj, Gandhinagar 382055, Gujarat, India. Email: Shweta Anand.

## Abstract

**Background:** Cognitive test performance in India is shaped by education, literacy and language as well as by neurodegeneration, which limits classification based on cognitive tests alone. We examined whether combining cognitive, informant-reported, sociodemographic and cardiometabolic information improves dementia classification, and which domains carry the gain.

**Methods:** We analyzed the 2,528 LASI-DAD Wave 1 respondents with a clinical consensus Clinical Dementia Rating (CDR). Dementia was defined as CDR of 1 or above (192 cases, 7.6%). Predictors were eight cognitive measures, the Informant Questionnaire on Cognitive Decline in the Elderly (IQCODE), and age, sex and education.Ten cardiometabolic assays were evaluated in the 1,788 respondents with blood results. Five algorithms were compared using nested cross-validation (5 outer, 3 inner folds). We assessed discrimination, calibration and net benefit, tested each domain’s contribution with paired bootstrap differences in ROC-AUC, and examined predictors with permutation importance and SHAP.

**Results:** Logistic regression achieved a ROC-AUC of 0.953 (95% CI 0.937 to 0.966), with calibration intercept 0.00, calibration slope 1.11 and Brier score 0.037. Random forest, gradient boosting and XGBoost performed similarly (0.946 to 0.947). Cognitive measures alone gave a ROC-AUC of 0.899. Adding the IQCODE increased it by 0.052 (95% CI 0.038 to 0.067); adding sociodemographic variables changed it by +0.0020 (−0.00004 to 0.0040) and adding cardiometabolic assays by +0.0032 (−0.0004 to 0.0073). At a training-derived threshold, sensitivity was 0.875, specificity 0.902, positive predictive value 0.422 and negative predictive value 0.989. Observed dementia prevalence rose from 2.1% to 93.5% across five risk strata, and the model gave greater net benefit than referring all or none at every threshold from 0.01 to 0.50. Oversampling with SMOTE did not change discrimination (0.952) but raised mean predicted risk to 21.2% against an observed 7.6%.

**Conclusions:** An interpretable logistic regression model classified clinically rated dementia with high discrimination and good calibration. Its advantage over cognitive tests came almost entirely from informant-reported decline; sociodemographic characteristics and cardiometabolic biomarkers added negligible discrimination. The clinical reference standard incorporated the same informant and cognitive information, and external validation is required before the model is used.

## 1. INTRODUCTION

Dementia classification remains difficult in ageing populations where cognitive performance is shaped by more than neurodegenerative change. Educational attainment, literacy, language background and health status all influence how individuals perform on cognitive tests, which makes scores difficult to interpret across heterogeneous populations (Ardila, 2005; Manly, 2006; Stern, 2009). The problem is acute in low- and middle-income settings, where dependence on any single screening measure may contribute to both under recognition and misclassification of dementia (Prince et al., 2015; World Health Organization, 2021).

One response is to combine information from several domains. Informant questionnaires such as IQCODE ask a relative to compare a person’s current everyday functioning with their functioning ten years earlier, and so measure change against the individual’s own baseline rather than against a population norm (Jorm, 2004). Vascular, metabolic and inflammatory measures reflect processes associated with dementia risk (Whitmer et al., 2005; Walker et al., 2019; Livingston et al., 2020). A model combining these domains may classify dementia more accurately than cognitive tests alone, but that proposition needs to be tested domain by domain rather than assumed.

The Longitudinal Ageing Study in India (LASI) and its diagnostic sub-study LASI-DAD provide nationally representative data on late-life cognition with clinical consensus ratings of dementia (Arokiasamy et al., 2012; Lee et al., 2020a). National estimates from these data place dementia prevalence among adults aged 60 and over at 7.4% (Lee et al., 2023a). Previous machine learning work in LASI-DAD has shown that models trained on the information presented to clinical raters reproduce consensus diagnoses with a ROC-AUC above 0.90 (Jin et al., 2021), has used a semi-supervised model to extend dementia classification to respondents without a consensus rating (Jin et al., 2023), and has identified the IQCODE and the Hindi Mental State Examination (HMSE) as the most influential assessments in an explainable model (Maupin et al., 2025). Cardiometabolic and inflammatory markers have been examined in relation to cognitive function in the same cohort (Hu et al., 2020).

What these studies do not establish is how much each domain contributes beyond the others, whether cardiometabolic biomarkers add anything to cognitive and informant measures for classification, and whether the resulting risk estimates are calibrated well enough to support risk stratification. This study develops and internally validates a multidomain classification model in LASI-DAD against clinical consensus ratings, quantifies the incremental contribution of each domain with paired tests, and assesses calibration and net benefit alongside discrimination. It follows the TRIPOD+AI reporting guideline (Collins et al., 2024).

## 2. METHODS

### 2.1 Data source and participants

LASI is a nationally representative longitudinal survey of adults aged 45 years and older in India (Arokiasamy et al., 2012). LASI-DAD recruited a subsample of LASI respondents aged 60 and older across 18 states and union territories between 2017 and 2020, using a two-stage stratified design that oversampled respondents at higher risk of cognitive impairment, and administered the Harmonized Cognitive Assessment Protocol with modifications for the Indian context together with an informant interview (Lee et al., 2020a). Wave 1 comprises 4,096 respondents. Analyses used Harmonized LASI-DAD Version A.4 (Lee et al., 2025), linked by participant identifier to the LASI-DAD venous blood assay results.

The primary sample comprised all 2,528 Wave 1 respondents with a clinical consensus rating, the same sample used in previous work (Jin et al., 2021; Maupin et al., 2025). Clinical assessment was carried out for respondents interviewed in fieldwork phases 2 and 3; the release records 2,471 of the rated respondents in those phases and 57 in phase 1. The phase in which a respondent was interviewed depended on fieldwork logistics, primarily state of residence, and not on individual characteristics such as cognitive status (Maupin et al., 2025). The cardiometabolic subsample comprised the 1,788 consensus-rated respondents with venous blood assay results. The 740 consensus-rated respondents without assays had a higher dementia prevalence (9.6%) than those with assays (6.8%), so the subsample is not a random subset of the primary sample.

Survey weights were not applied because the aim was individual-level classification rather than prevalence estimation. The sampling indicator for elevated cognitive risk is a design variable and was not used as a predictor.

### 2.2 Outcome

Dementia status was determined by an online clinical consensus procedure using the Clinical Dementia Rating (Morris, 1993). Each case was assigned to three CDR-certified clinicians who rated it independently on the six CDR domains from standardized interview and test data; discrepancies were resolved by discussion among the raters and, where needed, at a moderated online consensus meeting (Lee et al., 2020b; Lee et al., 2023b). Dementia was defined as a final consensus CDR of 1 or above, consistent with previous work in this cohort (Lee et al., 2023a; Maupin et al., 2025). Respondents with CDR 0.5 were classified as not having dementia.

The raters reviewed the cognitive assessment, self-reported functional difficulties, depression and anxiety, the informant interview including the IQCODE, sociodemographic characteristics and health history (Maupin et al., 2025). Several predictors in this study were therefore inputs to the reference standard, a form of incorporation bias that tends to raise estimates of classification accuracy (Worster and Carpenter, 2008). The dementia indicators distributed with LASI-DAD as 20 imputation variables were not used as the outcome. In the data they equal the consensus rating where one exists and for respondents without a consensus rating they are the predictions of a published semi-supervised model rather than clinical ratings (Jin et al., 2023). They are used only in a sensitivity analysis reproducing the version 1 configuration.

### 2.3 Predictors

Cognitive performance was represented by eight measures: orientation to time and place (0 to 10), immediate word recall summed across three learning trials of a ten-word list (0 to 30), delayed word recall (0 to 10), immediate logical memory story recall (0 to 50), clock drawing (0 to 3), animal fluency, and factor-analytic composite scores for memory and executive function derived by the LASI-DAD team (Gross et al., 2020). Informant-reported decline was measured with the 16-item IQCODE, scored as the mean rating across the items the informant answered from 1 to 5 (Jorm, 2004; Lee et al., 2025).

Sociodemographic predictors were the participant’s age at the LASI-DAD interview, sex and completed years of education. The cardiometabolic domain comprised glycated haemoglobin, total, HDL and LDL cholesterol, triglycerides, albumin, vitamin B12, 25-hydroxy vitamin D, homocysteine and high-sensitivity C-reactive protein. The source variable for each predictor is listed in Supplementary Table S1.

The HMSE total score was not used as a predictor. The HMSE is not administered identically to all respondents. Two items, reading and obeying a written command and writing a sentence, are given only to respondents who report that they can read and write; respondents who cannot are instead asked to copy a gesture made by the interviewer and to say a sentence about their house. The total is therefore assembled from partly different tasks in different tasks in different literacy groups (Ganguli et al., 1995; Lee et al., 2025). In the primary sample 60.0% of respondents could not read or write, and the proportion was higher among those with dementia (87.5%). The orientation items, which are common to all respondents, were retained. The effect of adding the HMSE total was examined in a sensitivity analysis.

### 2.4 Missing Data

Harmonized LASI-DAD cognitive and informant variables incorporate the study’s own imputation of item-level missing values, identified by imputation flags in the release. All 12 primary predictors were therefore complete for all 2,528 participants. In the cardiometabolic subsample, missingness ranged from 0% to 7.2% (vitamin B12), and missing assay values were imputed with k-nearest neighbours imputation (k = 5) fitted within training folds only (Troyanskaya et al., 2001).

### 2.5 Model development and internal validation

Five algorithms were compared: L2-penalized logistic regression, random forest, gradient boosting, XGBoost and a support vector machine with a radial basis kernel (Breiman, 2001; Chen and Guestrin, 2016; Pedregosa et al., 2011). Performance was estimated within nested cross-validation across the full sample, using five stratified outer folds for evaluation and three stratified inner folds for hyperparameter tuning by ROC-AUC (Varma and Simon, 2006). Imputation, standardization and model fitting took place within training folds and performance is reported on the pooled out-of-fold predictions. Hyperparameter grids are given in the analysis code.

The classification threshold was selected in each outer fold by maximizing Youden’s Index on that fold’s training data and averaged across folds. Discrimination was summarized by ROC-AUC with 95% confidence intervals from 2,000 bootstrap resamples, and by average precision. Calibration was assessed with the calibration intercept, the calibration slope, the Brier score and a calibration plot by decile of predicted risk (Van Calster et al., 2019). Clinical utility was assessed with decision-curve analysis across threshold probabilities from 0.01 to 0.50 (Vickers and Elkin, 2006). Predicted probabilities from logistic regression were grouped into equal-width risk strata (0 to 0.2, 0.2 to 0.4, 0.4 to 0.6, 0.6 to 0.8 and 0.8 to 1.0).

Class imbalance was not corrected in the primary models. Oversampling methods such as SMOTE (Chawla et al., 2002) distort predicted probabilities without improving discrimination in logistic regression (van den Goorbergh et al., 2022), and risk stratification depends on calibrated probabilities. SMOTE applied within training folds was evaluated as a sensitivity analysis.

### 2.6 Contribution of each domain

Logistic regression models were fitted in the primary sample with cognitive measures alone, with the IQCODE added, and with sociodemographic variables added, the last being the primary model. In the cardiometabolic subsample, the primary predictors were compared with the primary predictors plus the ten assays. Because all models used identical folds, differences in ROC-AUC were estimated from the paired out-of-fold predictions with 95% confidence intervals from 2,000 paired bootstrap resamples.

### 2.7 Interpretation

Permutation importance was computed on each outer test fold as the mean decrease in ROC-AUC across 20 random permutations of each predictor and averaged across folds (Breiman, 2001). SHAP values were computed on each outer test fold for the fitted logistic regression (Lundberg and Lee, 2017). Adjusted odds ratios per standard deviation, with 95% confidence intervals, were estimated from an unpenalized logistic regression fitted to the full primary sample. The coefficients of the final penalized model are reported in Supplementary Table S5.

### 2.8 Sensitivity analyses

Four sensitivity analyses were performed with logistic regression: the primary model with SMOTE; the primary model with the HMSE total added; and two models reproducing the version 1 configuration (the 2,892 respondents with assays, the averaged imputation-file dementia indicator, 22 predictors and SMOTE), one with the informant demographic variables and one with the participant’s own.

### 2.9 Software and reproducibility

Analyses were conducted in Python using pandas, scikit-learn, imbalance-learn, XGBoost, statsmodels and SHAP. The random seed was fixed at 42.

## 3. RESULTS

### 3.1 Participants

The primary sample comprised 2,528 participants, of whom 192 (7.6%) had dementia (Table 1). Mean age was 69.5 (7.5) years, 52.4% were female, 50.6% had no formal education and 60.0% could not read or write. Participants with dementia were older, more often female, had fewer years of education and higher IQCODE scores and performed worse on every cognitive measure.

**Table 1.** Characteristics of the primary sample, by dementia status and of the cardiometabolic subsample. Dementia is a clinical consensus CDR of 1 or above.

| Characteristic | Primary sample | Dementia | No dementia | Cardiometabolic subsample |
|---|---|---|---|---|
| N | 2528 | 192 | 2336 | 1788 |
| Age, years, mean (SD) | 69.5 (7.5) | 75.2 (9.0) | 69.1 (7.2) | 69.3 (7.2) |
| Female, n (%) | 1324 (52.4) | 118 (61.5) | 1206 (51.6) | 917 (51.3) |
| Education, years, mean (SD) | 3.6 (4.6) | 1.3 (2.8) | 3.8 (4.7) | 3.6 (4.5) |
| No formal education, n (%) | 1280 (50.6) | 146 (76.0) | 1134 (48.5) | 915 (51.2) |
| Cannot read or write, n (%) | 1516 (60.0) | 168 (87.5) | 1348 (57.7) | 1077 (60.2) |
| Rural, n (%) | 1655 (65.5) | 142 (74.0) | 1513 (64.8) | 1237 (69.2) |
| IQCODE, mean (SD) | 3.51 (0.57) | 4.46 (0.49) | 3.43 (0.50) | 3.50 (0.57) |
| Orientation (0 to 10), mean (SD) | 7.6 (2.3) | 4.2 (2.3) | 7.9 (2.0) | 7.6 (2.2) |
| Word recall, delayed (0 to 10), mean (SD) | 3.1 (2.2) | 1.3 (1.7) | 3.2 (2.2) | 3.1 (2.2) |
| Dementia (CDR ≥ 1), n (%) | 192 (7.6) | 192 (100.0) | 0 (0.0) | 121 (6.8) |

### 3.2 Discrimination and calibration

Logistic regression achieved a ROC-AUC of 0.953 (95% CI 0.937 to 0.966) and an average precision of 0.716 (Table 2, Figure 1). Random forest (0.947), XGBoost (0.947) and gradient boosting (0.946) performed similarly with overlapping confidence intervals. The support vector machine performed worse (0.900). Logistic regression was retained as the primary model on the basis of equivalent discrimination and full interpretability, consistent with evidence that machine learning methods offer no systematic advantage over logistic regression for clinical prediction (Christodoulou et al., 2019; Rudin, 2019).

**Table 2.** Discrimination and calibration of the five algorithms, pooled out-of-fold predictions (N = 2,528, 192 cases).

| **Model** | **ROC-AUC (95% CI)** | **Average precision** | **Brier score** | **Calibration intercept** | **Calibration slope** |
|---|---|---|---|---|---|
| Logistic regression | 0.953 (0.937 to 0.966) | 0.716 | 0.037 | 0.00 | 1.11 |
| Random forest | 0.947 (0.928 to 0.962) | 0.705 | 0.038 | 0.00 | 1.20 |
| XGBoost | 0.947 (0.931 to 0.962) | 0.698 | 0.039 | 0.04 | 0.90 |
| Gradient boosting | 0.946 (0.926 to 0.963) | 0.696 | 0.039 | 0.00 | 0.91 |
| SVM | 0.900 (0.870 to 0.927) | 0.653 | 0.043 | -0.15 | 0.85 |

**Figure 1.**
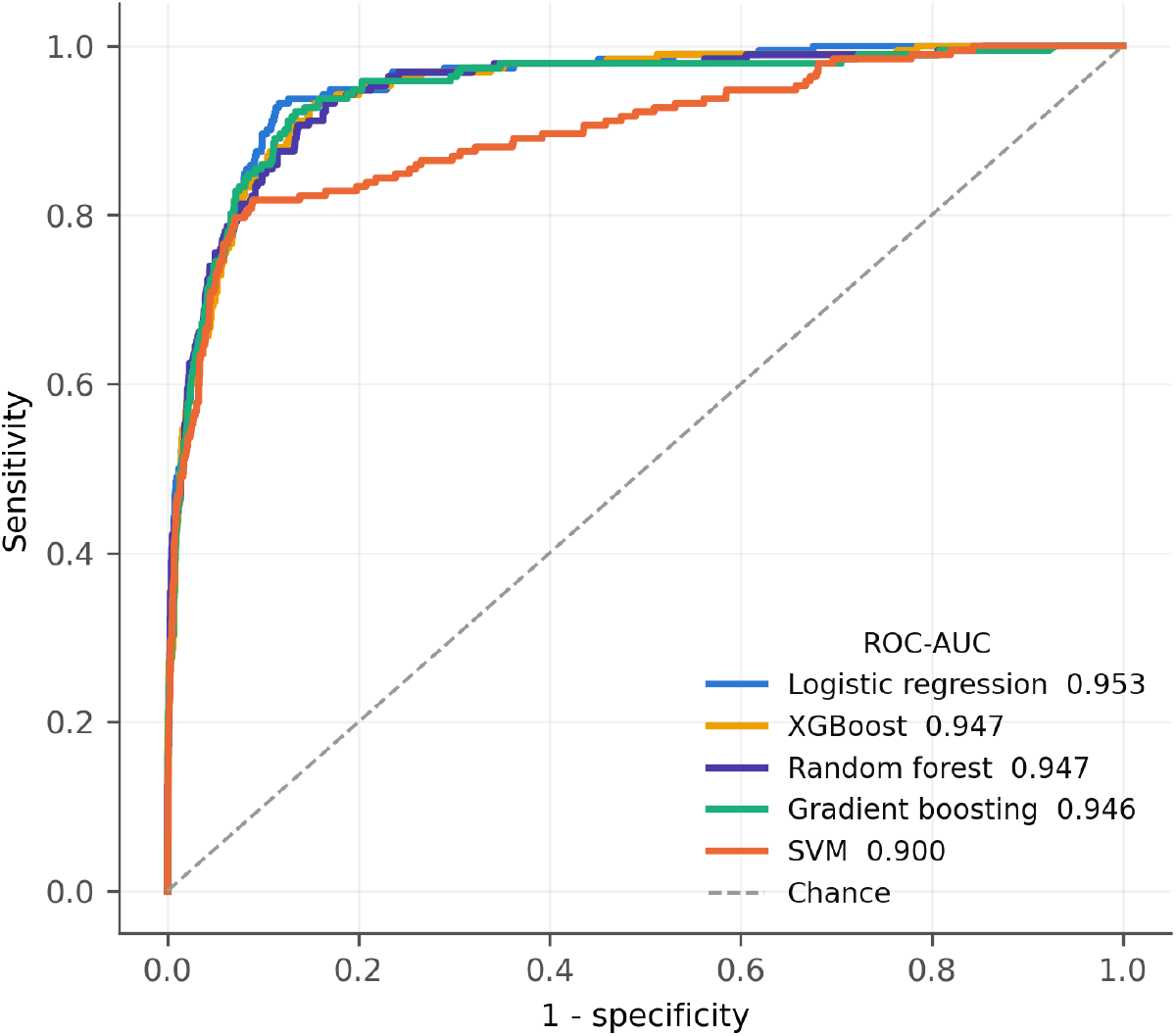
Receiver operating characteristic curves for the five algorithms, pooled out-of-fold predictions.

Logistic regression was well calibrated, with a calibration intercept of 0.00, a calibration slope of 1.11 and a mean predicted risk of 7.6% against an observed prevalence of 7.6% (Figure 2). The same model trained with SMOTE had essentially the same ROC-AUC (0.952) but a calibration intercept of −2.32, a mean predicted risk of 21.2% and a Brier score of 0.082, more than double that of the primary model.

**Figure 2.**
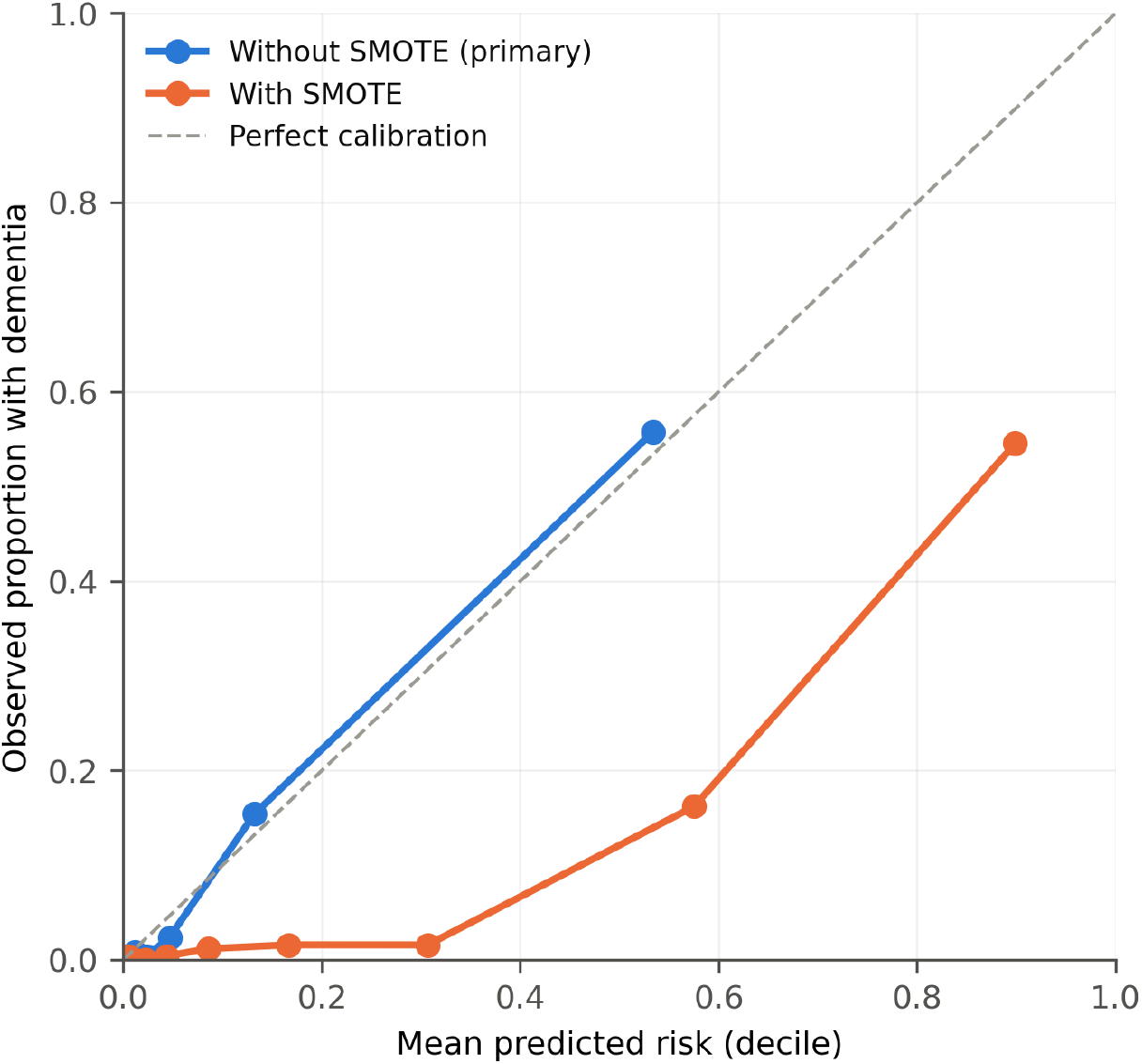
Calibration of logistic regression with and without SMOTE. Points are deciles of predicted risk. Most participants fall in the lowest deciles, so points cluster near the origin.

### 3.3 Contribution of each domain

Cognitive measures alone gave a ROC-AUC of 0.899 (95% CI 0.877 to 0.919). Adding the IQCODE raised it to 0.951, a paired difference of 0.052 (95% CI 0.038 to 0.067) and raised average precision from 0.489 to 0.713. Adding age, sex and education changed ROC-AUC by +0.0020 (95% CI −0.00004 to 0.0040). In the cardiometabolic subsample, the primary predictors gave a ROC-AUC of 0.949 and adding the ten assays changed it by +0.0032 (95% CI −0.0004 to 0.0073) (Table 3). Of the total gain of 0.054 over cognitive measures in the primary sample, 96.3% came from the IQCODE.

**Table 3.**
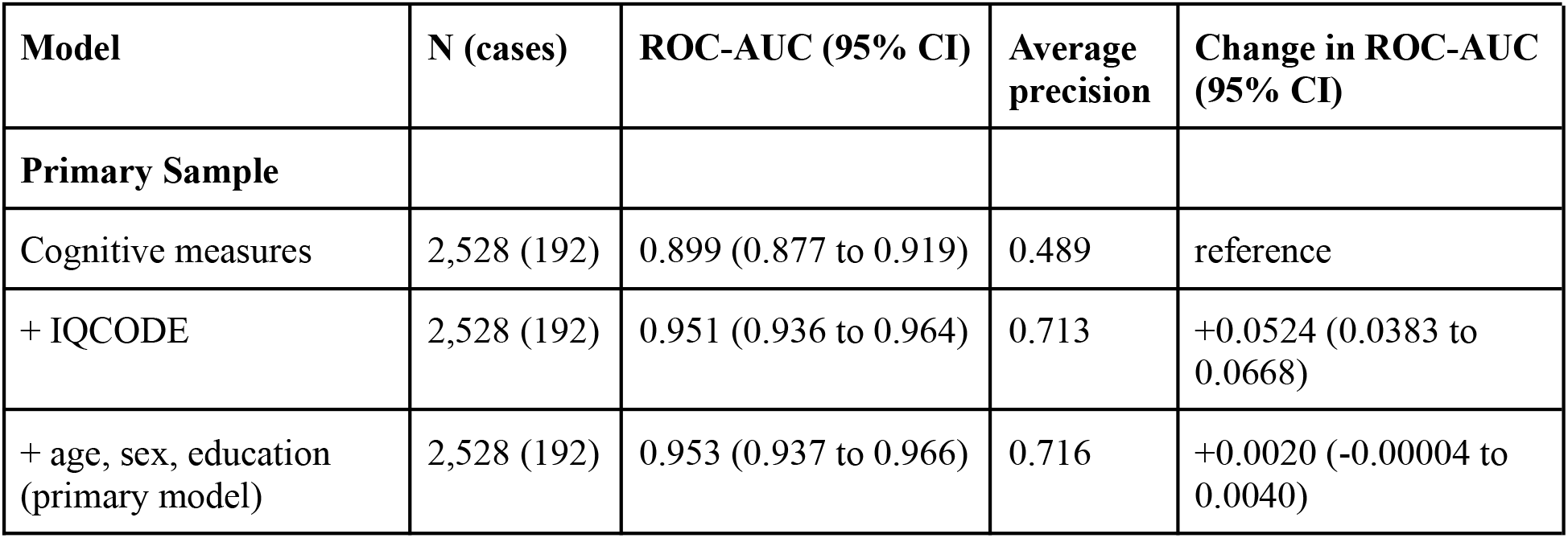
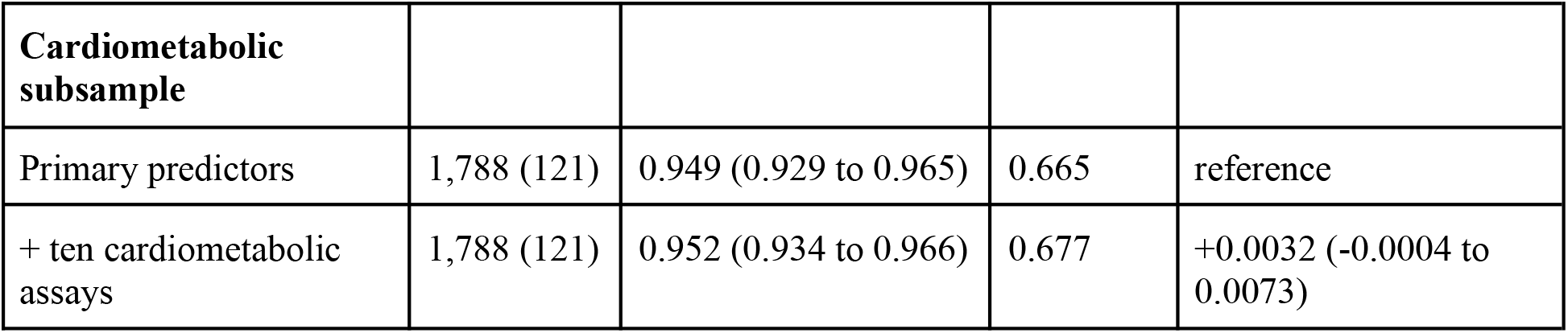
Incremental contribution of each domain, logistic regression. Each change is relative to the row above within the same sample, from paired bootstrap resampling of out-of-fold predictions.

| **Model** | **N (cases)** | **ROC-AUC (95% CI)** | **Average precision** | **Change in ROC-AUC (95% CI)** |
|---|---|---|---|---|
| **Primary Sample** |  |  |  |  |
| Cognitive measures | 2,528 (192) | 0.899 (0.877 to 0.919) | 0.489 | reference |
| + IQCODE | 2,528 (192) | 0.951 (0.936 to 0.964) | 0.713 | +0.0524 (0.0383 to 0.0668) |
| + age, sex, education (primary model) | 2,528 (192) | 0.953 (0.937 to 0.966) | 0.716 | +0.0020 (-0.00004 to 0.0040) |
| **Cardiometabolic subsample** |  |  |  |  |
| Primary predictors | 1,788 (121) | 0.949 (0.929 to 0.965) | 0.665 | reference |
| + ten cardiometabolic assays | 1,788 (121) | 0.952 (0.934 to 0.966) | 0.677 | +0.0032 (-0.0004 to 0.0073) |

### 3.4 Operating characteristics and clinical utility

At the training-derived threshold of 0.117, sensitivity was 0.875, specificity 0.902, positive predictive value 0.422 and negative predictive value 0.989. The model identified 168 of 192 cases with 230 false positives, 1.4 false positives per case detected and missed 24 cases (Table 4).

**Table 4.** Operating characteristics of the primary logistic regression model.

| **Measure** | **Value** |
|---|---|
| Threshold (training-derived) | 0.117 |
| Sensitivity | 0.875 |
| Specificity | 0.902 |
| Positive predictive value | 0.422 |
| Negative predictive value | 0.989 |
| True positives / false positives | 168 / 230 |
| False negatives / true negatives | 24 / 2106 |

In decision-curve analysis, the model gave greater net benefit than referring all participants or none at every threshold probability from 0.01 to 0.50 (Figure 3). At a threshold of 0.10, net benefit was 0.058 for the model against −0.027 for referring everyone.

**Figure 3.**
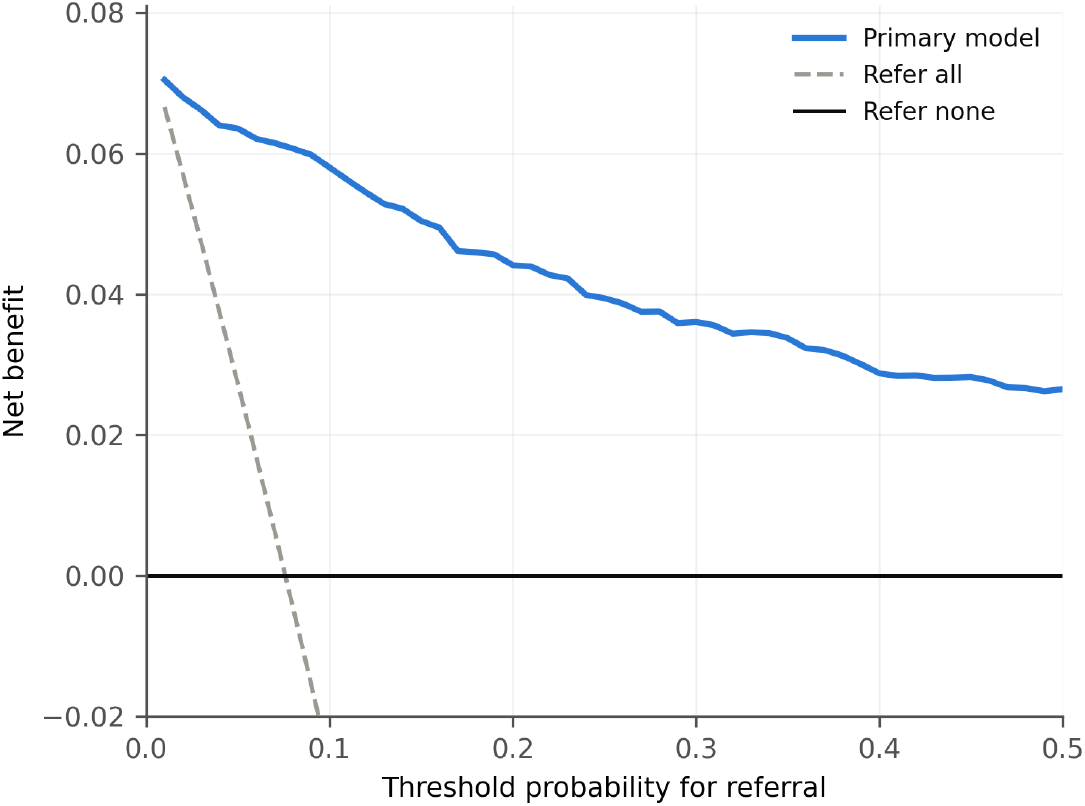
Decision-curve analysis for the primary model. Net benefit is plotted against the threshold probability at which referral would be made.

Observed dementia prevalence increased across the five risk strata from 2.1% in the very low stratum to 93.5% in the very high stratum (Figure 4, Supplementary Table S2). The two highest strata contained 102 participants (4.0% of the sample) and 85 of the 192 cases (44.3%). The very low stratum contained 89.2% of participants and 48 cases, 25.0% of all cases.

**Figure 4.**
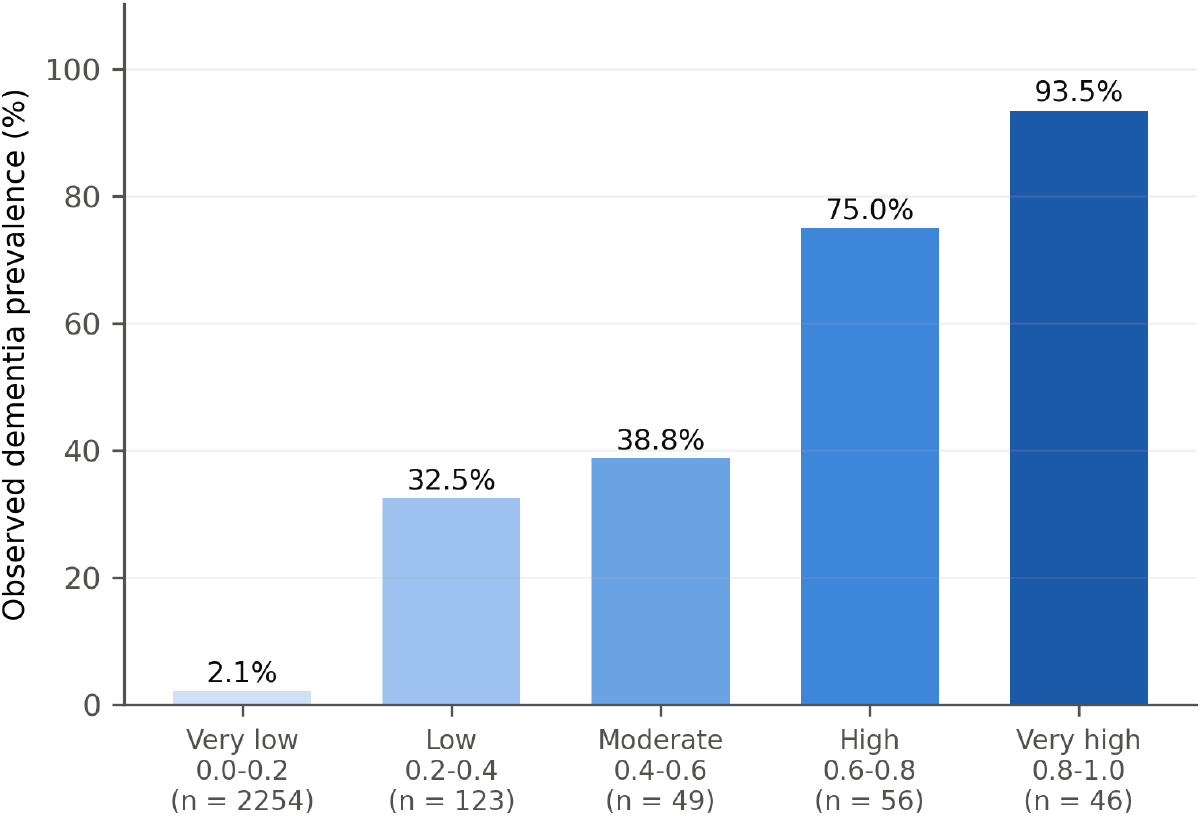
Observed dementia prevalence across equal-width strata of predicted risk from the primary model.

### 3.5 Predictor contributions

The IQCODE had the largest permutation importance, a mean decrease in ROC-AUC of 0.129, followed by orientation (0.042) and the executive composite (0.011). No other predictor reduced ROC-AUC by more than 0.002 when permuted (Figure 5). SHAP values showed the same ordering: higher IQCODE scores increased predicted risk, and better orientation and executive performance decreased it (Figure 6).

**Figure 5.**
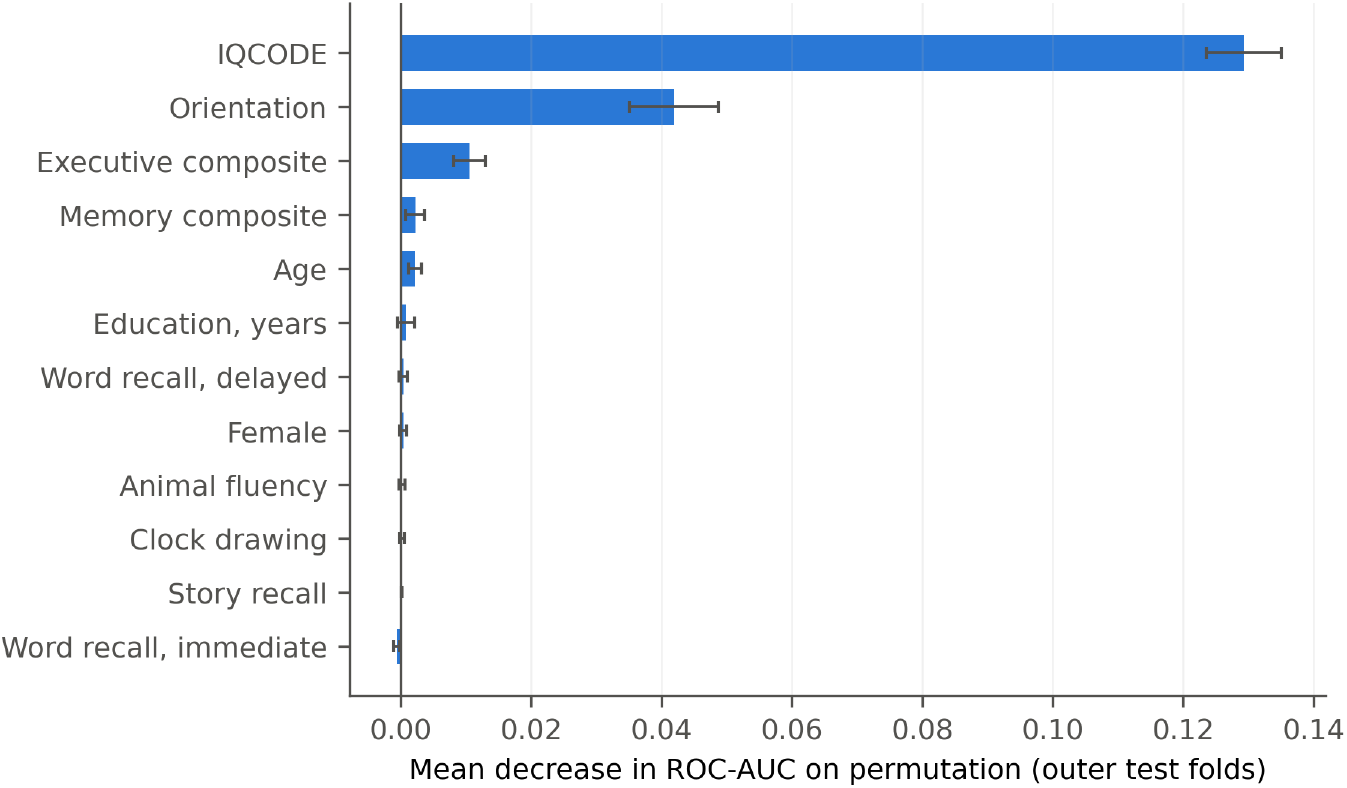
Permutation importance, mean decrease in ROC-AUC on the outer test folds. Error bars show the standard deviation across folds.

**Figure 6.**
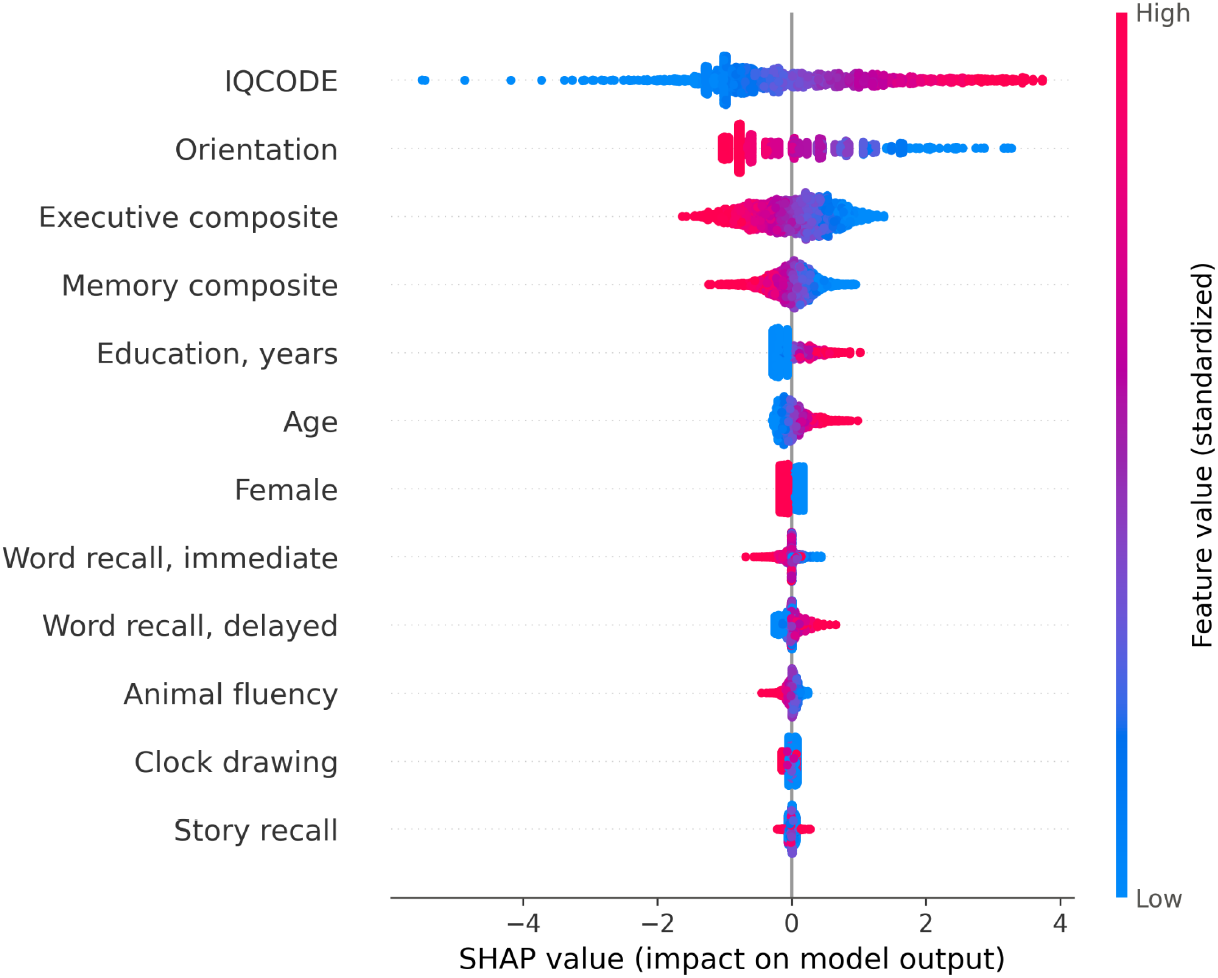
SHAP values for the primary logistic regression on the outer test folds. Each point is one participant; colour shows the standardized predictor value.

In the unpenalized model, the adjusted odds ratio per standard deviation was 4.77 (95%CI 3.73 to 6.08) for the IQCODE, 0.34 (95% CI 0.25 to 0.46) for orientation and 0.43 (95% CI 0.27 to 0.70) for the executive composite (Supplementary Table S3). Older age (odds ratio 1.22, 95% CI 1.01 to 1.47) and more years of education (odds ratio 1.60, 95% CI 1.08 to 2.38) were also associated with higher odds. Word recall, story recall, clock drawing, animal fluency, the memory composite and sex were not independently associated with dementia in this model (all 95% confidence intervals included 1) which is expected given the strong correlation among the cognitive measures.

### 3.6 Sensitivity analyses

Adding the HMSE total to the primary model left ROC-AUC unchanged at 0.953 (Supplementary Table S4).

## 4 DISCUSSION

In 2,528 LASI-DAD respondents with a clinical consensus rating, a logistic regression model combining cognitive measures, informant-reported decline and sociodemographic characteristics classified dementia with a ROC-AUC of 0.953 and was well calibrated. The multidomain gain over cognitive tests was real but came almost entirely from one domain. The IQCODE added 0.052 to ROC-AUC, while sociodemographic characteristics and ten cardiometabolic assays each added less than 0.005. More flexible algorithms did not improve on logistic regression.

These results agree with previous work in this cohort. Models trained on the information available to clinical raters reproduce consensus diagnoses with a ROC-AUC above 0.90 (Jin et al., 2021) and an explainable XGBoost model identified the IQCODE and the HMSE as the two most influential assessments (Maupin et al., 2025). The present analysis adds a direct, paired estimate of what each domain contributes and shows that a model of 12 predictors, calibrated and interpretable, reaches the same level of discrimination.

The strength of the IQCODE is consistent with its design. It asks whether a person has changed relative to their own earlier functioning which makes it less dependent on schooling than tests that compare performance with a population norm (Jorm, 2004). In a sample in which half of participants had no formal education, that property is likely to matter. Two cautions apply. The clinical raters had access to the IQCODE and the cognitive test results, so the reference standard is not independent of the predictors and some of the IQCODE’s apparent value reflects incorporation bias (Worster and Carpenter, 2008). The finding also does not establish that informant report can replace cognitive testing because both were present in the model.

The absence of a contribution from cardiometabolic assays should be read narrowly. These markers were measured in late life, at the same time as the outcome, and the question here is whether they improve classification of established dementia given detailed cognitive and informant information, not whether they are associated with dementia risk over the life course (Whitmer et al., 2005; Walker et al., 2019; Livingston et al., 2020). Within the resolution of this analysis they did not improve classification. This reverses the version 1 conclusion that cardiometabolic variables provided consistent incremental contributions.

Education was positively associated with dementia once cognitive performance was held constant. At a given test score, a more educated person was more likely to have been rated as having dementia. This is consistent with cognitive reserve and with education-related differences in test performance, whereby the same score represents a larger departure from premorbid functioning in a person with more schooling (Stern, 2009; Ardila, 2005). The estimate is conditional on the other predictors and should not be interpreted as a causal effect of education.

Calibration matters for how the model could be used. The risk strata rest on the predicted probabilities being close to observed risk. Oversampling with SMOTE, used in version 1, left discrimination unchanged but produced a mean predicted risk nearly three times the observed prevalence, which would have misrepresented risk in every stratum (van den Goorbergh et al., 2022). Without oversampling, predicted risk agreed closely with observed risk overall, with slight overestimation in the lower-middle deciles (4.6% predicted against 2.4% observed in the eighth decile) and the model gave positive net benefit at all threshold probabilities examined. Its limits are also clear. At the selected threshold 230 if 398 positive results (57.8%) were false and 25.0% of cases fell in the lowest risk stratum, so the model would not remove the need for clinical assessment.

## 4.1 Limitations

The model was validated internally only. Performance in other populations, other survey waves and clinical settings is unknown and external validation is required.

The predictors overlap with the information used by the clinical raters which is expected to inflate estimated accuracy. A reference standard independent of the predictors, such as longitudinal progression or biomarker-defined pathology, would address this.

Respondents with CDR 0.5 were grouped with those without dementia. The model therefore distinguishes dementia from everything else and does not address the detection of milder impairment.

The cardiometabolic subsample had a lower dementia prevalence than consensus-rated respondents without assays (6.8 % against 9.6%), so the null result for the assays applies to a selected group.

Survey weights were not applied and the results describe the analytic sample rather than the population of older adults in India.

The harmonized cognitive and informant variables include the study’s imputation of item-level missing values, and the analysis is always cross-sectional.

The eight cognitive predictors come from an extended research battery administered by trained interviewers. The model is not a brief screening instrument.

### 4.2 Next steps

External validation in independent cohorts is the priority. LASI-DAD Wave 2 allows the model to be tested against incident dementia at follow-up which would also provide a reference standard that does not incorporate the Wave 1 predictors. A model restricted to the IQCODE and a small number of cognitive items, evaluated for calibration across education and literacy groups, would test whether a much shorter assessment can achieve similar performance.

## 5. CONCLUSIONS

A calibrated and interpretable logistic regression model classified clinically rated dementia in a nationally representative Indian cohort with high discrimination. Its advantage over cognitive tests came almost entirely from informant-reported decline. Sociodemographic characteristics and cardiometabolic biomarkers added negligible discrimination. Because the clinical reference standard incorporated the same information and the model has been validated only internally, it should be regarded as a basis for external validation rather than a tool ready for use.

## Data Availability

All data produced in the present work are contained in the manuscript.

## ACKNOWLEDGEMENTS

Data were obtained from the Harmonized Diagnostic Assessment of Dementia for the Longitudinal Aging Study in India (LASI-DAD), produced and distributed by the University of Southern California with funding from the National Institute on Aging (R01AG051125, U01AG064948). We are grateful to the LASI-DAD team and to the study participants.

## ETHICS

LASI-DAD received ethical approval from the University of Southern California Institutional Review Board (UP15-00684) and the Indian Council of Medical Research for the All India Institute of Medical Sciences (54/01/Indo-foreign/Ger/16-NCD-II) (Lee et al., 2023b). This study is a secondary analysis of deidentified data obtained through the data provider’s registration process and involved no contact with participants.

## DATA AND CODE AVAILABILITY

Data from LASI and LASI-DAD are available to registered researchers through the Longitudinal Ageing Study in India data repository at the International Institute for Population Sciences (https://www.iipsindia.ac.in) for the former and through the Gateway to Global Aging Data (https://g2aging.org) and the Alzheimer’s Disease Data Initiative AD workbench for the latter.

## SUPPLEMENTARY MATERIALS

**Supplementary Table S1.** Source variables for each predictor.

| **Predictor** | **Variable used** |
|---|---|
| Orientation (0 to 10) | r1orient_t5 + r1orient_p5 |
| Word recall, immediate (0 to 30) | r1word_total |
| Word recall, delayed (0 to 10) | r1word_d |
| Story recall (0 to 50) | r1mb_imm |
| Clock drawing (0 to 3) | r1dr_clock3 |
| Animal fluency | r1verbal |
| Memory composite | r1bmemory |
| Executive composite | r1bexefu |
| IQCODE (1 to 5) | r1jormscore |
| Age | r1hagey (participant) |
| Sex | ragender (participant) |
| Education | raedyrs (participant) |
| Glycated haemoglobin (HbA1c) | hba1c__glycated_haemogl2 |
| Total cholesterol | cholesterol__total2 |
| HDL cholesterol | hdl_cholesterol2 |
| LDL cholesterol | ldl_cholesterol2 |
| Triglycerides | triglycerides_level2 |
| Albumin | albumin2 |
| Vitamin B12 | vitamin_b12_level2 |
| 25-hydroxy Vitamin D | *25*hydroxy__oh__vit_d2 |
| Homocysteine | homocysteine2 |
| hsCRP | hscrphigh_sensitivity_c2 |
| Outcome | r1cdr_final ≥ 1 (clinical consensus) |

*Variables prefixed r1 or ra are from Harmonized LASI-DAD Version A.4; assay and dm\_variables are from the merged LASI-DAD release. This study uses the LASI-DAD factor-analytic broad-domain scores for memory and executive function.*

**Supplementary Table S2.**
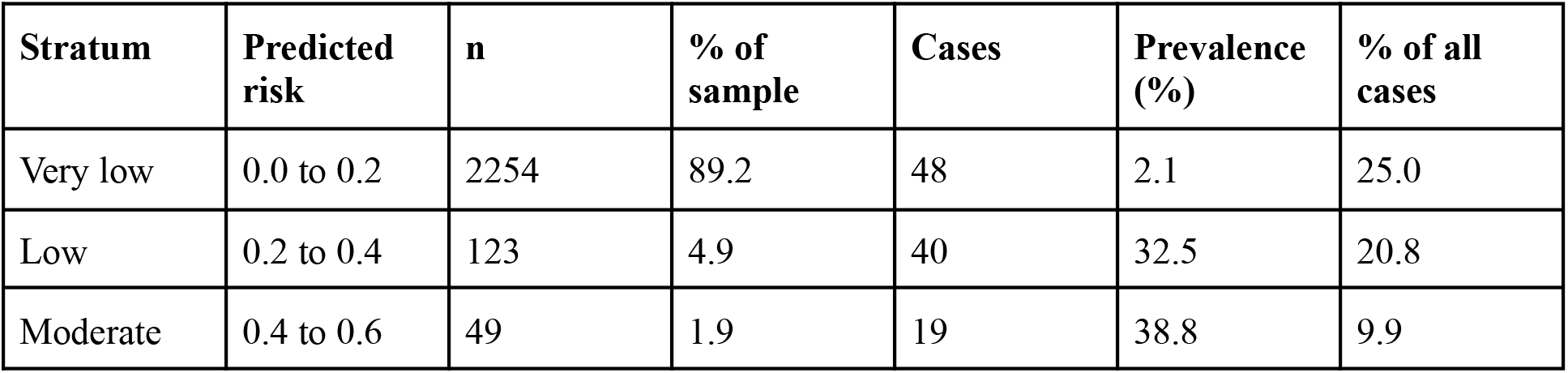
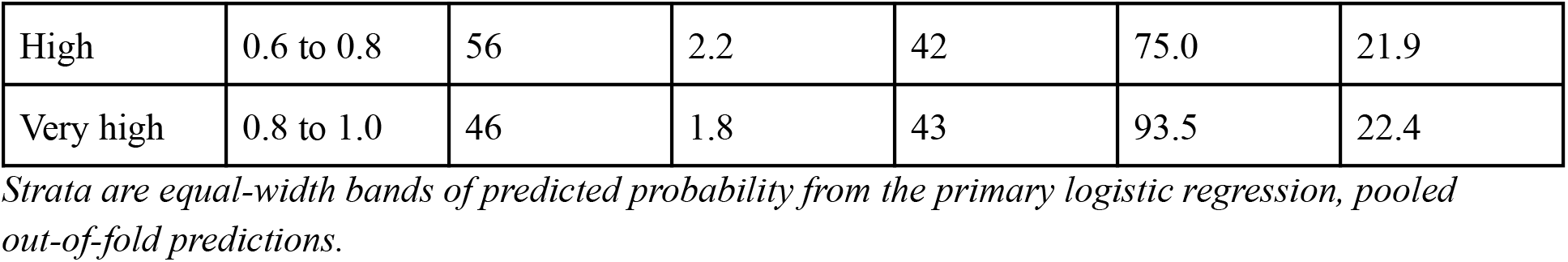
Observed dementia prevalence across risk strata.

| **Stratum** | **Predicted risk** | **n** | **% of sample** | **Cases** | **Prevalence (%)** | **% of all cases** |
|---|---|---|---|---|---|---|
| Very low | 0.0 to 0.2 | 2254 | 89.2 | 48 | 2.1 | 25.0 |
| Low | 0.2 to 0.4 | 123 | 4.9 | 40 | 32.5 | 20.8 |
| Moderate | 0.4 to 0.6 | 49 | 1.9 | 19 | 38.8 | 9.9 |
| High | 0.6 to 0.8 | 56 | 2.2 | 42 | 75.0 | 21.9 |
| Very high | 0.8 to 1.0 | 46 | 1.8 | 43 | 93.5 | 22.4 |

*Strata are equal-width bands of predicted probability from the primary logistic regression, pooled out-of-fold predictions.*

**Supplementary Table S3.** Adjusted odds ratios per standard deviation.

| **Predictor** | **Odds ratio per SD** | **95% CI** | **p** |
|---|---|---|---|
| Orientation | 0.34 | 0.25 to 0.46 | <0.001 |
| Word recall, immediate | 0.99 | 0.68 to 1.42 | 0.942 |
| Word recall, delayed | 1.24 | 0.87 to 1.79 | 0.236 |
| Story recall | 1.13 | 0.72 to 1.76 | 0.607 |
| Clock drawing | 1.02 | 0.68 to 1.53 | 0.910 |
| Animal fluency | 1.03 | 0.78 to 1.34 | 0.855 |
| Memory composite | 0.65 | 0.33 to 1.27 | 0.203 |
| Executive composite | 0.43 | 0.27 to 0.70 | <0.001 |
| IQCODE | 4.77 | 3.73 to 6.08 | <0.001 |
| Age | 1.22 | 1.01 to 1.47 | 0.036 |
| Female | 0.84 | 0.66 to 1.07 | 0.163 |
| Education, years | 1.60 | 1.08 to 2.38 | 0.020 |

*Unpenalized logistic regression on the full primary sample (N = 2528, 192 cases), all 12 predictors entered together. For Female the odds ratio is per SD of a binary indicator.*

**Supplementary Table S4.**
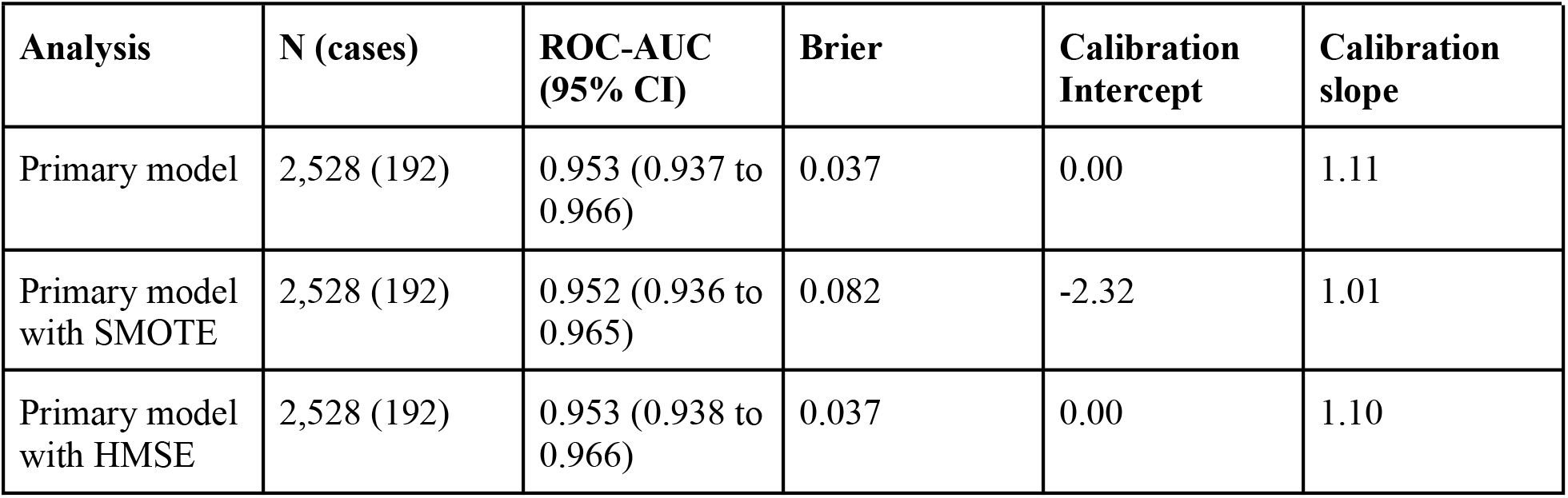
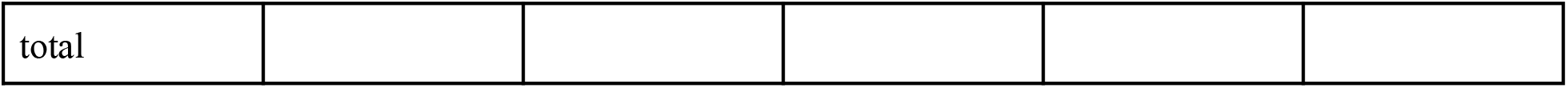
Sensitivity analyses, logistic regression.

| **Analysis** | **N (cases)** | **ROC-AUC (95% CI)** | **Brier** | **Calibration Intercept** | **Calibration slope** |
|---|---|---|---|---|---|
| Primary model | 2,528 (192) | 0.953 (0.937 to 0.966) | 0.037 | 0.00 | 1.11 |
| Primary model with SMOTE | 2,528 (192) | 0.952 (0.936 to 0.965) | 0.082 | -2.32 | 1.01 |
| Primary model with HMSE | 2,528 (192) | 0.953 (0.938 to 0.966) | 0.037 | 0.00 | 1.10 |
| total |  |  |  |  |  |

**Supplementary Table S5.** Final model.

| **Predictor** | **Mean** | **SD** | **Coefficient per SD** |
|---|---|---|---|
| Orientation | 7.573 | 2.284 | -0.9062 |
| Word recall, immediate | 11.615 | 4.932 | -0.0672 |
| Word recall, delayed | 3.087 | 2.246 | 0.1097 |
| Story recall | 8.389 | 7.739 | 0.0124 |
| Clock drawing | 1.071 | 1.081 | -0.0384 |
| Animal fluency | 10.941 | 4.495 | -0.0628 |
| Memory composite | -0.015 | 0.890 | -0.2903 |
| Executive composite | -0.049 | 0.905 | -0.5907 |
| IQCODE | 3.512 | 0.569 | 1.3492 |
| Age | 69.522 | 7.498 | 0.1977 |
| Female | 0.524 | 0.499 | -0.1247 |
| Education, years | 3.642 | 4.602 | 0.2598 |
| Intercept |  |  | -4.5814 |

*L2-penalized logistic regression fitted to the full primary sample (C = 0.1). Linear predictor = intercept + sum of coefficient x (value - mean) / SD; predicted risk = 1 / (1+exp (-linear predictor)). Female is coded 1 for women and 0 for men.*

**Supplementary Figure S1.**
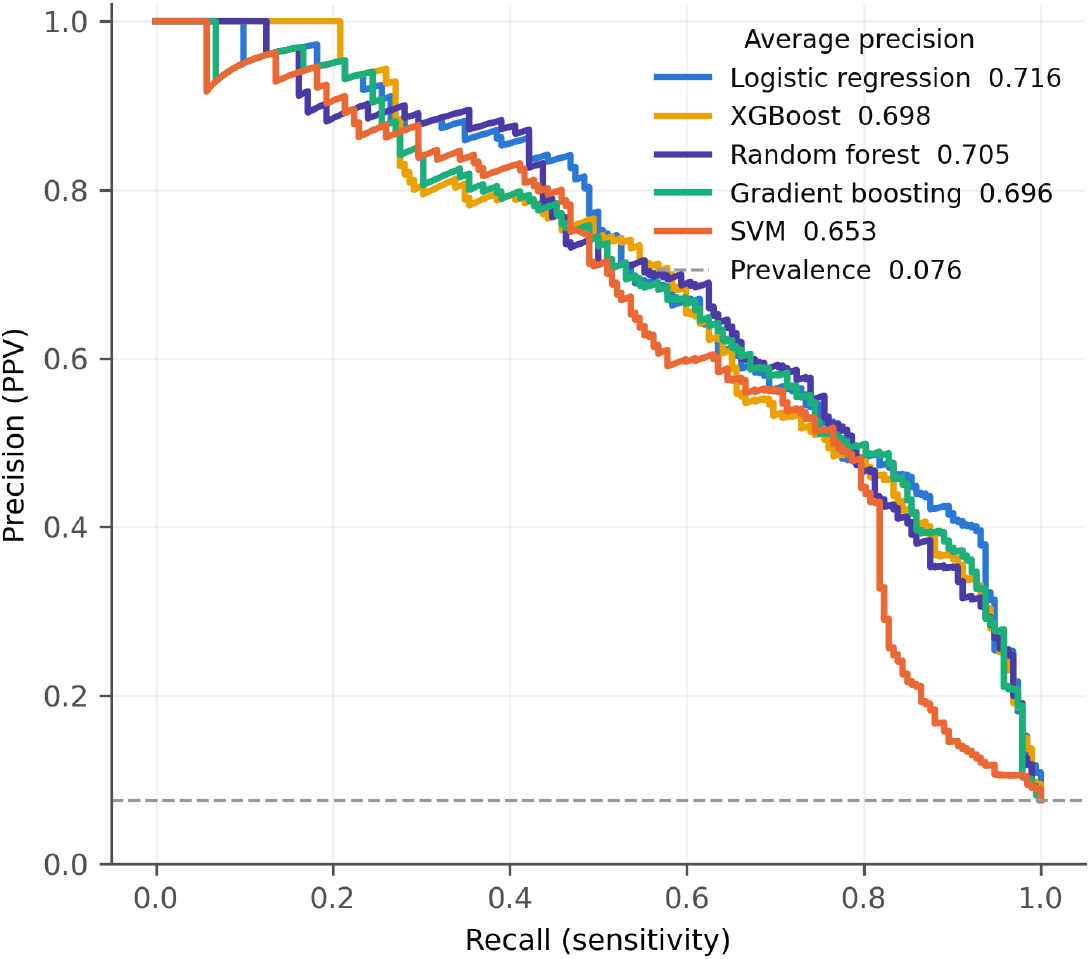
Precision-recall curves for the five algorithms, pooled out-of-fold predictions. The dashed line is the sample prevalence.

